# Employment status, occupational profile, and common mental disorders among workers in urban informal settlements in Brazil

**DOI:** 10.64898/2026.04.01.26350007

**Authors:** Julia Fernandes Cavalcanti Prestes, Thayane Silva Nunes, Fábio Neves Souza, Diogo César de Carvalho Santiago, Yeimi Alzate López, Fabiana Almerinda Gonçalves Palma, Juliet Oliveira Santana, Priscilla Elizabeth Ferreira dos Santos, Daiana de Olieveira, Adedayo Michael Awoniyi, Christine E. Stauber, Federico Costa, Cleber Cremonese

## Abstract

Urban informal settlements (referred to as favelas in Brazil), reflect longstanding socioeconomic and racial inequalities and are home to a workforce frequently exposed to precarious employment conditions. This study describes the socio-occupational characteristic and estimates the prevalence of common mental disorders (CMDs) among workers residing in five urban informal communities in Salvador, Bahia, Brazil. A cross-sectional epidemiological study (n=587) was conducted with formal and informal workers aged 18-70 years. The outcome was measured using the Self-Reporting Questionnaire-SRQ-20, and associations were evaluated using Poisson Regression, with analysis stratified by employment type. Data analysis was performed using R 3.6.0+ software. The overall prevalence of CMD was 14.0%, increasing to 22.7% among informal workers. In the adjusted analysis of the overall sample, informal employment and persistent fear of job loss were associated with a higher prevalence of mental health problems, whereas the 40-49 age groups showed a lower prevalence of CMD compared with younger workers. In stratified analyses, female sex and job insecurity were associated with CMD among formal workers, while lower monthly income (<$181) was an important among informal workers. This pioneering study highlights the role of precarious employment conditions in the social determination of mental health distress among residents of urban informal communities.

## INTRODUCTION

Common mental disorders (CMDs) are characterized by non-psychotic symptoms typically associated with anxiety and depressive conditions, including insomnia, difficulty concentrating, depressed mood, irritability, feelings of worthlessness, anhedonia, recurrent crying episodes, suicidal ideation, and impaired work functioning [1,2]. According to the World Health Organization (WHO) - World Mental Health Report: Transforming Mental Health for All, the global prevalence of mental disorders reached approximately 970 million people in 2019, representing about 13% of the world’s population [3]. The burden of these conditions is disproportionately concentrated in low- and middle-income countries (LMICs), such as Brazil which account for an estimated 82% of cases. In Brazil, nationally representative studies examining population mental health remain limited. Systematic national surveillance began only in 2013, when the Brazilian National Health Survey (Pesquisa Nacional de Saúde-PNS) first incorporated mental health indicators. Data from the 2019 survey indicated that 10.2% of Brazilians reported having received a diagnosis of depression at some point in their lives [4].

CMDs are shaped by a complex interplay of individual, family and community, and broader structural factors. These relationships are dynamic and multifaceted, making precise casual pathways difficult to disentangle [3]. Employment type constitutes a key determinant of CMD risk, with numerous studies showing greater vulnerability among workers engaged in precarious or informal employment [4,5–8]. According to the International Labor Organization [9], precarious employment is characterized by both objective and subjective form of insecurity. Objective insecurities include limited labor rights, unstable contractual arrangements, and low wages, whereas subjective insecurities encompass uncertainty, job-related stress, work overload, and feelings of devaluation associated with certain types of employment.

Precarious work can occur in both formal and informal employment types. In the Brazilian context, informal employment is defined by the absence of legally guaranteed labor protections established under the Consolidation of Labor Laws (Consolidação das Leis do Trabalho-CLT). Workers in informal employment typically lack access to key labor rights, including paid sick leave, paid vacation, a 13th-month salary, and contributions to social security. As a result, they are often exposed to greater employment instability, income insecurity, workplace exploitation, and occupational hazards [10–13]. Informal employment remains widespread across Latin America and the Caribbean. Recent estimates indicate that approximately 53.7% of employed workers in the region are engaged in informal work, while 19.8 million individuals are unemployed, representing 6.1% of the workforce [14]. In Brazil, 37.8% of the employed workforce was engaged in informal sector in the second quarter of 2025, while the unemployment rate reached 5.8% of the economically active population (EAP) [15].

However, the workforce conditions are unevenly distributed across Brazil. Informality and unemployment are disproportionately concentrated in the North and Northeast regions, areas that historically have larger populations of Black and Indigenous communities. In this context, Bahia, the Brazilian state with the highest proportion of Black residents [16], is particularly affected. In the second quarter of 2025, Bahia recorded the second-highest unemployment rate in the country (9.1%), nearly double the national average, and an informality rate of 52.3%, the third highest among Brazilian states [15]. The strong overlap between race and socioeconomic vulnerability reflects the enduring influence of structural racism on Brazil and Bahia workforce, shaped by historical legacies of colonialism and slavery [17].

Urban informal settlements (often referred to as favelas in Brazil) are a prominent feature of cities shaped by deep social and racial inequality [18,19]. These territories have historically emerged through processes of self-settlement, as marginalized populations seek access to housing and livelihood opportunities within larger urban centers [20–23]. According to UN-Habitat [24], approximately 12.5% of the global population of around 1.1 billion people resides in informal settlements. In Brazil, an estimated 16 million people (8.1% of the population) live in such settings IBGE [20]. In Salvador, the capital of Bahia, approximately 34.9% of residents reside in urban communities, representing the third-highest proportion among Brazilian capitals [20].

In these communities, multiple factors are often associated with mental health risks [25–27]. Among these, employment types play a critical role, as individuals frequently engage in precarious or informal work to sustain their livelihoods. Despite these vulnerabilities, epidemiological studies examining the occupational and mental conditions of workers residing in urban informal settlements in Brazil remain scarce. This study therefore aims to describe the socio-occupational profile of workers and estimate the prevalence of CMDs by employment type among workers living in urban informal settlements in the city of Salvador, Bahia, between 2023 and 2025.

## METHODS

### Study design and research context

From December 2023 to March 2025, a cross-sectional epidemiological study nested within a longitudinal survey was conducted in six urban informal settlements in Salvador. The larger parent study evaluates the impact of health interventions aimed at preventing urban leptospirosis and other health outcomes between 2020 and 2026 [28]. Among the observational studies nested within the longitudinal survey was an investigation assessing the prevalence of CMDs among residents of the surveyed populations. The present study is therefore a cross-sectional epidemiological investigation conducted in five of the six urban informal settlements included in the larger project, as the final community was recruited only after completion of this study.

### Study population and sample size

According to the Demographic Census (2022) [20], Salvador contains 262 urban informal settlements, with approximately 1,033,258 residents, representing 42.7% of the city’s population. The parent cohort study determined its sample size using an independent calculation [28]. Given the present study examined a different outcome, a new sample size calculation was performed using the EpiInfo™ version 7.2. Assuming a 20% prevalence of CMDs [29], a 95% confidence level, a 5% margin of error, and 90% statistical power, the required sample size was estimated at approximately 584 participants. Eligible participants were residents aged 18-70 years who slept at home at least three nights per week and had engaged in any economic activity in the previous month, regardless of employment type or job characteristics. Individuals with physical or mental health conditions that prevented completion of the questionnaires were excluded.

### Data collection procedure

Data were collected through household interviews conducted between December 2023 and March 2025. Two structured questionnaires were administered: the first captured participants’ demographic, socioeconomic, and occupational characteristics, and the second evaluated CMDs using Self-Reporting Questionnaire (SRQ-20) [30]. A team of 12 trained technicians conducted the interviews using standardized procedures. Data were recorded electronically in RedCap software, using smartphones [31].

### Data collection and analysis of study outcome

CMDs were evaluated using the SRQ-20, a 20-item screening instrument that assesses non-psychotic symptoms experienced in the past 30 days. Responses were recorded dichotomously (yes, no). The cutoff point used to indicate probable cases were ≥6 affirmative responses for males (89% sensitivity and 81% specificity), and ≥8 for females (86% sensitivity and 77% specificity) [30].

### Collection and analysis of sociodemographic variables

In order to analyze the sociodemographic profile, the following variables were chosen: sex (male, female); age group (18-29, 30-39, 40-49, 50-59, 60-70); race/color (white, black, brown, yellow/indigenous); educational attainment, i.e. years of study (≥12, 9-11, 0-8); marital status (married/stable union, single, widowed/divorced); main caregiver of a child (no, yes); and for those with at least one living child, it was asked if they were a beneficiary of Bolsa Família (yes, no).

### Collection and analysis of occupational variables

To characterize the occupational profile, several variables were examined, encompassing both objective aspects of employment relationships and subjective occupational characteristics. The variable "form of employment relationship" was originally collected in 12 categories: 1) worker governed by CLT; 2) formal fixed-term contract; 3) informal employee; 4) informal self-employed worker; 5) informal daily service provider; 6) self-employed professional; 7) formal employer; 8) informal employer; 9) internship/scholarship; 10) young apprentice; 11) civil servant; and 12) others. Due to low observations in many categories, analyses focused on the four most frequent categories: namely CLT, informal self-employed worker, informal employee, and others. For this recategorization, seven employment types (formal fixed-term contract, self-employed professional, formal employer, informal employer, internship/scholarship, young apprentice, and civil servant) were grouped under “others”. Additionally, the category “informal daily service provider” was merged with “informal employee”, as both represents informal labor arrangements involving a contracting party or employer. Based on the variable employment type, another variable-type of employment relationship, was created to classify participants as formal workers (CLT, civil servants, self-employed professionals, formal employers, formal fixed-term contract workers, and young apprentices), informal workers (informal employees, informal self-employed workers, informal daily service providers, and informal employers), or others (internship/scholarship and other unspecified categories). The variable “current occupation” was initially collected using the following categories: bricklayer, urban sanitation worker (garbage collector), street vendor, shop or fixed-business salesperson, recycler, domestic worker, cleaner (day laborer), general services worker, doorman/concierge, security guard, teacher, cook, administrative assistant, nanny, receptionist, driver, other (with an open-ended response-see supplementary material 1), and don’t know/no response. Responses were subsequently reviewed and grouped into six most frequent occupational categories: 1) construction and maintenance, 2) commerce, services, and food, 3) security, transport, logistics and urban sanitation, 4) domestic services, 5) administrative services, and 6) other.

Additional variables were included to further characterize participant’s occupational profile. These comprised: engagement in “casual work” in the past month (no, yes), daily working hours (>8h, 8h, 6h-7h, ≤ 5h); time in the main occupation (> 3 years, ≤ 3years), and monthly labor income, categorized into quartiles ≥$327, $256-326, $181-255, <$181. Additional variables included work location (Same neighborhood, nearby neighborhood, distant neighborhood, or another municipality) and the primary settings of workplace (private company, public institution, streets/squares/parks or other public spaces, own residence, or another person’s residence).

To evaluate social and subjective dimensions of labor relations among study population, participants responded to items measured on a Likert-type frequency scale, which were collapsed into three categories for analysis. The variables included fear of losing one’s job, perceived freedom to perform work activities, and emotional demands at work, each categorized as always/almost always, sometimes/rarely, or never.

### Data processing and analysis

Sociodemographic and occupational characteristics, as well as the prevalence of CMDs, were described using absolute and relative frequencies and stratified by type of employment. The distribution of CMDs across independent variables was summarized using prevalence estimates (Tables 1 and 2). Association between independent variables and the outcome were examined using the Chi-square test or Fisher’s exact test, as appropriate (Tables 3 and 4). Given the complexity of the outcome, variables with theoretical relevance and those with p-value ≤ 0.10 in bivariate analyses were considered for inclusion in the final models. Crude prevalence ratios (cPR) and adjusted prevalence ratios (aPR), with 95% confidence intervals, were estimated using Poisson regression model with robust variance [32,33]. All analyses were conducted using R software (version 3.6.0^+^). Although some stratified groups had relatively small numbers of observations, the analyses were retained to provide descriptive insight into this understudied population and to explore possible patterns across groups. Further studies are needed to confirm these hypotheses.

**Table 1.**
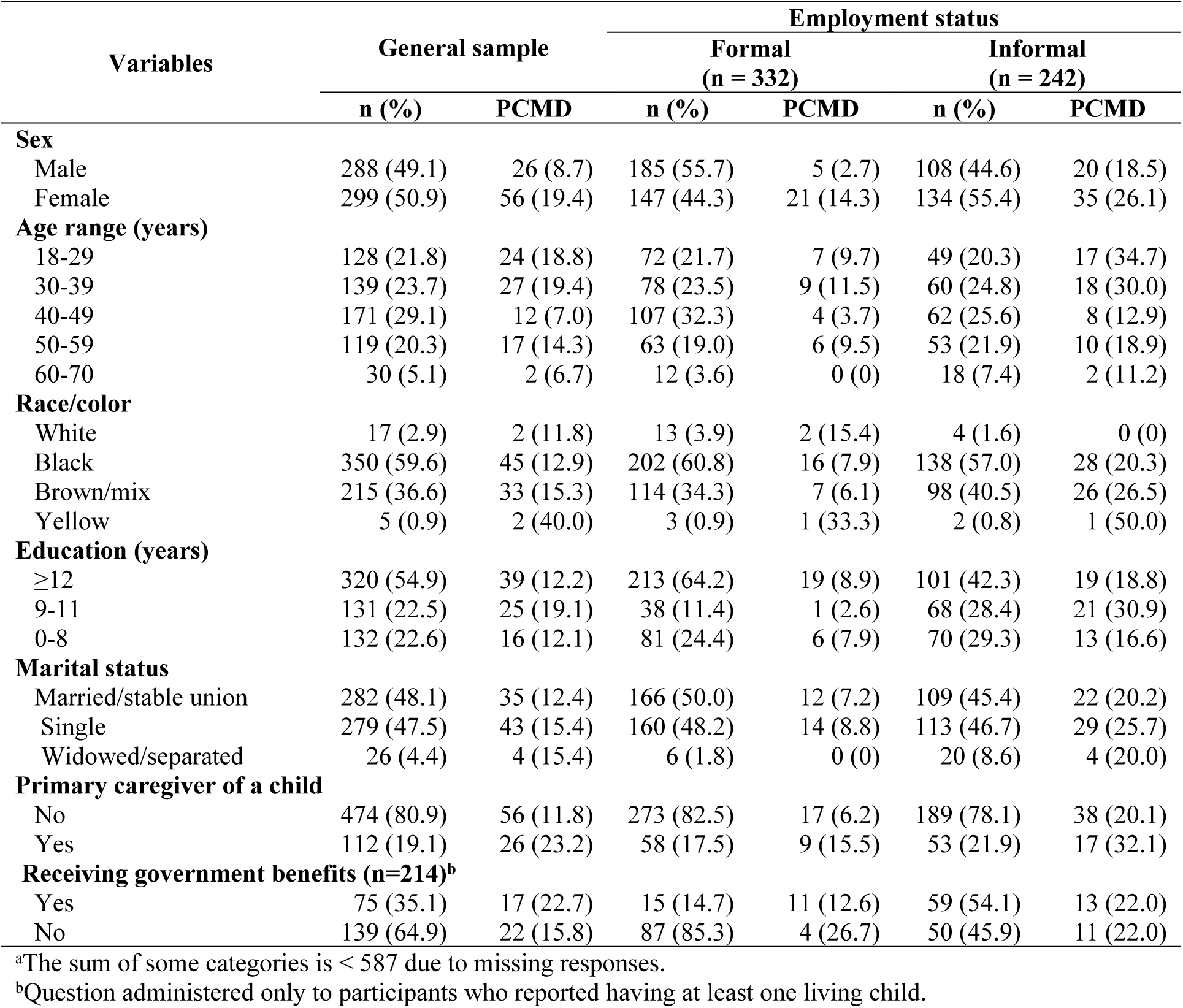
Distribution of sociodemographic characteristics and prevalence of common mental disorders (PCMD) among workers from urban informal settlements in Salvador, Bahia, Brazil, 2023-2025, by overall sample and employment status (n = 587)^a^

**Table 2.**
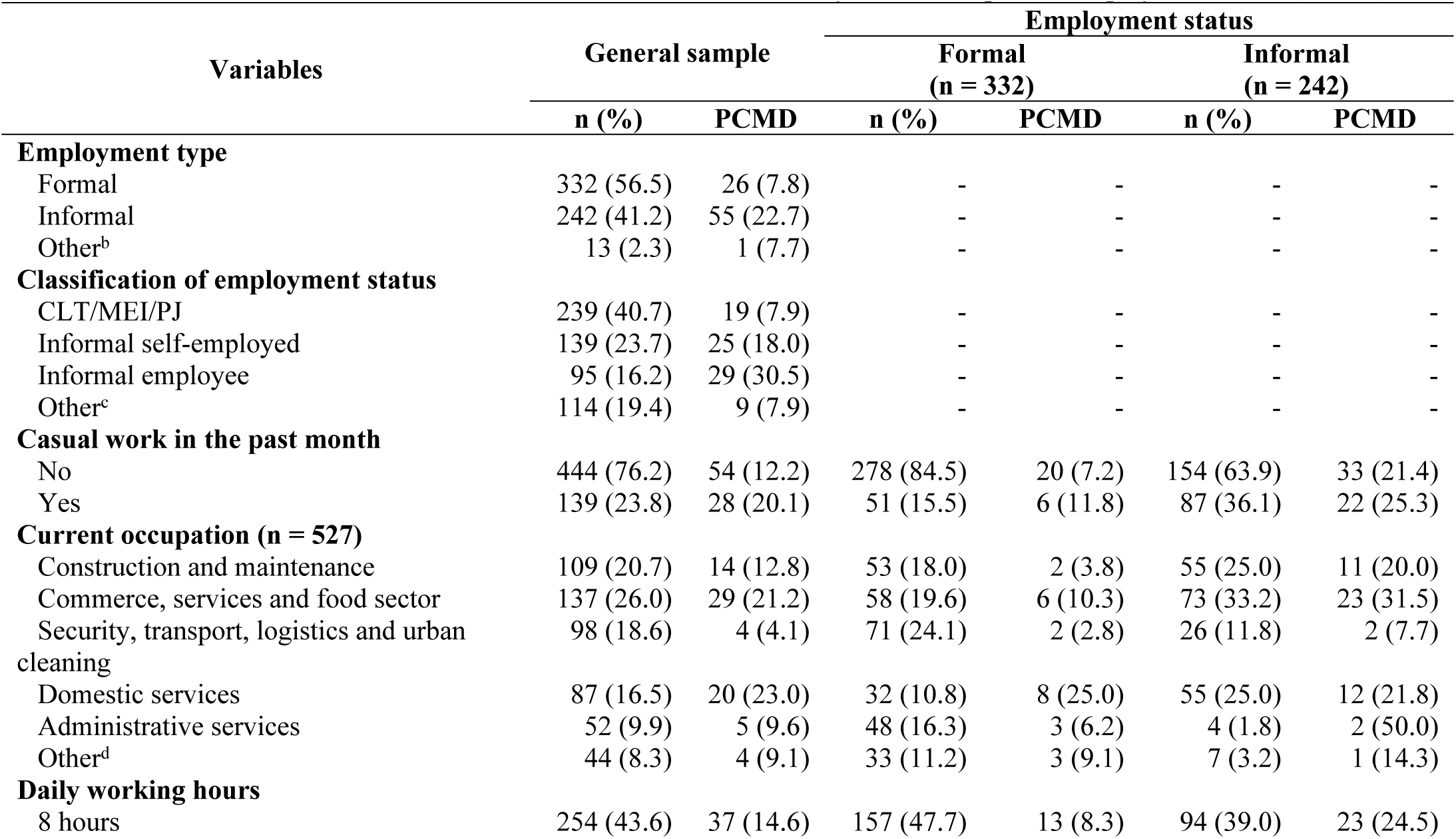
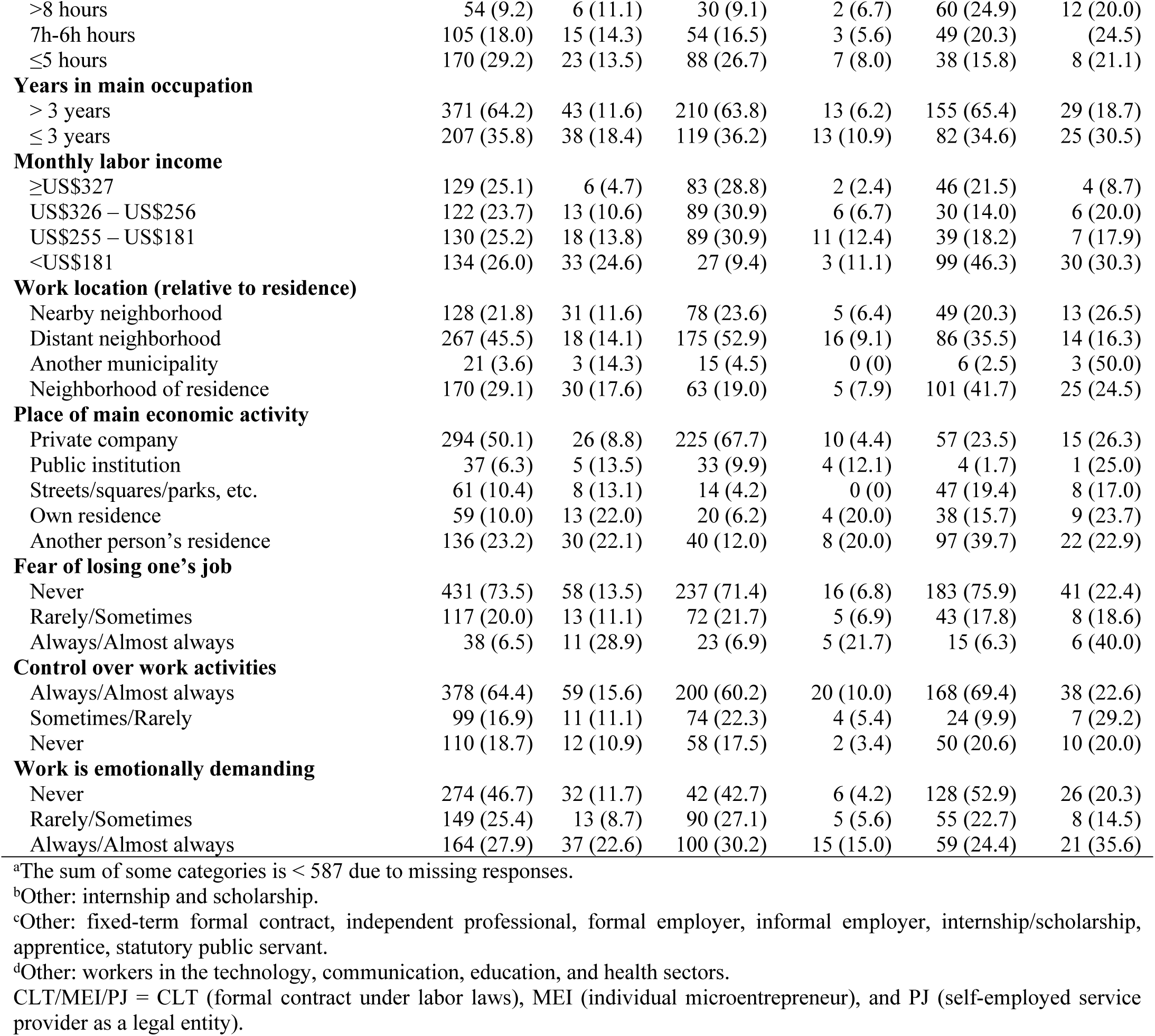
Distribution of occupational characteristics and prevalence of common mental disorders (PCMD) among workers from urban informal settlements in Salvador, Bahia, Brazil, 2023–2025, by overall sample and employment status (n = 587)^a^

| Variables | General sample |  | Employment status |  |  |  |
| --- | --- | --- | --- | --- | --- | --- |
|  |  |  | Formal<br>(n = 332) |  | Informal<br>(n = 242) |  |
|  | n (%) | PCMD | n (%) | PCMD | n (%) | PCMD |
| **Employment type** |  |  |  |  |  |  |
| Formal | 332 (56.5) | 26 (7.8) | - | - | - | - |
| Informal | 242 (41.2) | 55 (22.7) | - | - | - | - |
| Other<sup>b</sup> | 13 (2.3) | 1 (7.7) | - | - | - | - |
| **Classification of employment status** |  |  |  |  |  |  |
| CLT/MEI/PJ | 239 (40.7) | 19 (7.9) | - | - | - | - |
| Informal self-employed | 139 (23.7) | 25 (18.0) | - | - | - | - |
| Informal employee | 95 (16.2) | 29 (30.5) | - | - | - | - |
| Other<sup>c</sup> | 114 (19.4) | 9 (7.9) | - | - | - | - |
| **Casual work in the past month** |  |  |  |  |  |  |
| No | 444 (76.2) | 54 (12.2) | 278 (84.5) | 20 (7.2) | 154 (63.9) | 33 (21.4) |
| Yes | 139 (23.8) | 28 (20.1) | 51 (15.5) | 6 (11.8) | 87 (36.1) | 22 (25.3) |
| **Current occupation (n = 527)** |  |  |  |  |  |  |
| Construction and maintenance | 109 (20.7) | 14 (12.8) | 53 (18.0) | 2 (3.8) | 55 (25.0) | 11 (20.0) |
| Commerce, services and food sector | 137 (26.0) | 29 (21.2) | 58 (19.6) | 6 (10.3) | 73 (33.2) | 23 (31.5) |
| Security, transport, logistics and urban cleaning | 98 (18.6) | 4 (4.1) | 71 (24.1) | 2 (2.8) | 26 (11.8) | 2 (7.7) |
| Domestic services | 87 (16.5) | 20 (23.0) | 32 (10.8) | 8 (25.0) | 55 (25.0) | 12 (21.8) |
| Administrative services | 52 (9.9) | 5 (9.6) | 48 (16.3) | 3 (6.2) | 4 (1.8) | 2 (50.0) |
| Other<sup>d</sup> | 44 (8.3) | 4 (9.1) | 33 (11.2) | 3 (9.1) | 7 (3.2) | 1 (14.3) |
| **Daily working hours** |  |  |  |  |  |  |
| 8 hours | 254 (43.6) | 37 (14.6) | 157 (47.7) | 13 (8.3) | 94 (39.0) | 23 (24.5) |
| >8 hours | 54 (9.2) | 6 (11.1) | 30 (9.1) | 2 (6.7) | 60 (24.9) | 12 (20.0) |
| 7h-6h hours | 105 (18.0) | 15 (14.3) | 54 (16.5) | 3 (5.6) | 49 (20.3) | (24.5) |
| ≤5 hours | 170 (29.2) | 23 (13.5) | 88 (26.7) | 7 (8.0) | 38 (15.8) | 8 (21.1) |
| **Years in main occupation** |  |  |  |  |  |  |
| > 3 years | 371 (64.2) | 43 (11.6) | 210 (63.8) | 13 (6.2) | 155 (65.4) | 29 (18.7) |
| ≤ 3 years | 207 (35.8) | 38 (18.4) | 119 (36.2) | 13 (10.9) | 82 (34.6) | 25 (30.5) |
| **Monthly labor income** |  |  |  |  |  |  |
| ≥US\$327 | 129 (25.1) | 6 (4.7) | 83 (28.8) | 2 (2.4) | 46 (21.5) | 4 (8.7) |
| US\$326 – US\$256 | 122 (23.7) | 13 (10.6) | 89 (30.9) | 6 (6.7) | 30 (14.0) | 6 (20.0) |
| US\$255 – US\$181 | 130 (25.2) | 18 (13.8) | 89 (30.9) | 11 (12.4) | 39 (18.2) | 7 (17.9) |
| <US\$181 | 134 (26.0) | 33 (24.6) | 27 (9.4) | 3 (11.1) | 99 (46.3) | 30 (30.3) |
| **Work location (relative to residence)** |  |  |  |  |  |  |
| Nearby neighborhood | 128 (21.8) | 31 (11.6) | 78 (23.6) | 5 (6.4) | 49 (20.3) | 13 (26.5) |
| Distant neighborhood | 267 (45.5) | 18 (14.1) | 175 (52.9) | 16 (9.1) | 86 (35.5) | 14 (16.3) |
| Another municipality | 21 (3.6) | 3 (14.3) | 15 (4.5) | 0 (0) | 6 (2.5) | 3 (50.0) |
| Neighborhood of residence | 170 (29.1) | 30 (17.6) | 63 (19.0) | 5 (7.9) | 101 (41.7) | 25 (24.5) |
| **Place of main economic activity** |  |  |  |  |  |  |
| Private company | 294 (50.1) | 26 (8.8) | 225 (67.7) | 10 (4.4) | 57 (23.5) | 15 (26.3) |
| Public institution | 37 (6.3) | 5 (13.5) | 33 (9.9) | 4 (12.1) | 4 (1.7) | 1 (25.0) |
| Streets/squares/parks, etc. | 61 (10.4) | 8 (13.1) | 14 (4.2) | 0 (0) | 47 (19.4) | 8 (17.0) |
| Own residence | 59 (10.0) | 13 (22.0) | 20 (6.2) | 4 (20.0) | 38 (15.7) | 9 (23.7) |
| Another person's residence | 136 (23.2) | 30 (22.1) | 40 (12.0) | 8 (20.0) | 97 (39.7) | 22 (22.9) |
| **Fear of losing one's job** |  |  |  |  |  |  |
| Never | 431 (73.5) | 58 (13.5) | 237 (71.4) | 16 (6.8) | 183 (75.9) | 41 (22.4) |
| Rarely/Sometimes | 117 (20.0) | 13 (11.1) | 72 (21.7) | 5 (6.9) | 43 (17.8) | 8 (18.6) |
| Always/Almost always | 38 (6.5) | 11 (28.9) | 23 (6.9) | 5 (21.7) | 15 (6.3) | 6 (40.0) |
| **Control over work activities** |  |  |  |  |  |  |
| Always/Almost always | 378 (64.4) | 59 (15.6) | 200 (60.2) | 20 (10.0) | 168 (69.4) | 38 (22.6) |
| Sometimes/Rarely | 99 (16.9) | 11 (11.1) | 74 (22.3) | 4 (5.4) | 24 (9.9) | 7 (29.2) |
| Never | 110 (18.7) | 12 (10.9) | 58 (17.5) | 2 (3.4) | 50 (20.6) | 10 (20.0) |
| **Work is emotionally demanding** |  |  |  |  |  |  |
| Never | 274 (46.7) | 32 (11.7) | 42 (42.7) | 6 (4.2) | 128 (52.9) | 26 (20.3) |
| Rarely/Sometimes | 149 (25.4) | 13 (8.7) | 90 (27.1) | 5 (5.6) | 55 (22.7) | 8 (14.5) |
| Always/Almost always | 164 (27.9) | 37 (22.6) | 100 (30.2) | 15 (15.0) | 59 (24.4) | 21 (35.6) |

<sup>a</sup>The sum of some categories is < 587 due to missing responses.
<sup>b</sup>Other: internship and scholarship.
<sup>c</sup>Other: fixed-term formal contract, independent professional, formal employer, informal employer, internship/scholarship, apprentice, statutory public servant.
<sup>d</sup>Other: workers in the technology, communication, education, and health sectors.
CLT/MEI/PJ = CLT (formal contract under labor laws), MEI (individual microentrepreneur), and PJ (self-employed service provider as a legal entity).

**Table 3.** Crude prevalence ratios (cPR) for common mental disorders (CMD) and associated sociodemographic characteristics among workers from urban informal settlements in Salvador, Bahia, Brazil, 2023–2025, by overall sample and employment status.

| Variables | General sample |  | Employment status |  |  |  |
| --- | --- | --- | --- | --- | --- | --- |
|  | cPR (CI95%) | p-value | Formal |  | Informal |  |
|  |  |  | cPR (CI95%) | p-value | cPR (CI95%) | p-value |
| **Sex** |  | **<0.001** |  | **<0.001** |  | **<0.001** |
| Male | 1.00 |  | 1.00 |  | 1.00 |  |
| Female | **2.24 (1.42–3.62)** |  | **5.29 (2.15–15.83)** |  | 1.41 (0.82–2.49) |  |
| **Age range (years)** |  | **0.004** |  | 0.228 |  | **0.028** |
| 18-29 | 1.00 |  | 1.00 |  | 1.00 |  |
| 30-39 | 1.04 (0.60–1.81) |  | 1.19 (0.44–3.19) |  | 0.86 (0.44–1.69) |  |
| 40-49 | **0.37 (0.18–0.73)** |  | 0.38 (0.11–1.31) |  | **0.37 (0.15–0.84)** |  |
| 50-59 | 0.76 (0.40–1.41) |  | 0.98 (0.33–2.91) |  | 0.54 (0.24–1.17) |  |
| 60-70 | 0.36 (0.06–1.20) |  | 0.00 (0.00–Inf) |  | 0.32 (0.05–1.12) |  |
| **Race/color** |  | 0.284 |  | 0.152 |  |  |
| White | 1.00 |  | 1.00 |  | - |  |
| Black | 1.09 (0.34–6.70) |  | 0.51 (0.15–3.26) |  | - |  |
| Brown/mix | 1.30 (0.40–8.05) |  | 0.40 (0.10–2.68) |  | - |  |
| Yellow | 3.40 (0.41–28.34) |  | 2.17 (0.10–22.62) |  | *** |  |
| **Education (years)** |  | 0.128 |  | 0.528 |  | 0.130 |
| ≥12 | 1.00 |  | 1.00 |  | 1.00 |  |
| 9-11 | 1.57 (0.94–2.57) |  | 0.30 (0.02–1.42) |  | 1.77 (0.89–3.48) |  |
| 0-8 | 0.99 (0.54–1.75) |  | 0.83 (0.30–1.96) |  | 1.09 (0.57–2.06) |  |
| **Marital status** |  | 0.534 |  | 0.586 |  | 0.586 |
| Married/stable union | 1.00 |  | - |  | 1.00 |  |
| Single | 1.24 (0.80–1.95) |  | - |  | 1.27 (0.73–2.24) |  |
| Widowed/separated | 1.24 (0.37–3.10) |  | *** |  | 0.99 (0.29–2.59) |  |
| **Primary caregiver of a child** |  | **0.002** |  | **0.028** |  | **0.028** |
| No | 1.00 |  | 1.00 |  | 1.00 |  |
| Yes | **1.96 (1.22–3.09)** |  | **2.49 (1.06–5.47)** |  | **1.60 (0.88–2.78)** |  |
| **Receiving government benefits** |  | 0.293 |  | 0.435 |  | 1 |
| Yes | 1.00 |  | 1.00 |  | 1.00 |  |
| No | 0.70 (0.37–1.33) |  | 0.47 (0.16–1.71) |  | 1.00 (0.44–2.23) |  |

p-value: Chi-square test or Fisher test for heterogeneity–statistical significance. Values in bold indicate variables included in the multivariable analysis.
\*\*\*It was not possible to estimate this variable in the bivariate model due to the presence of values very close to zero.
CI, confidence interval 95%.
Value 1.00 = reference category.

**Table 4.**
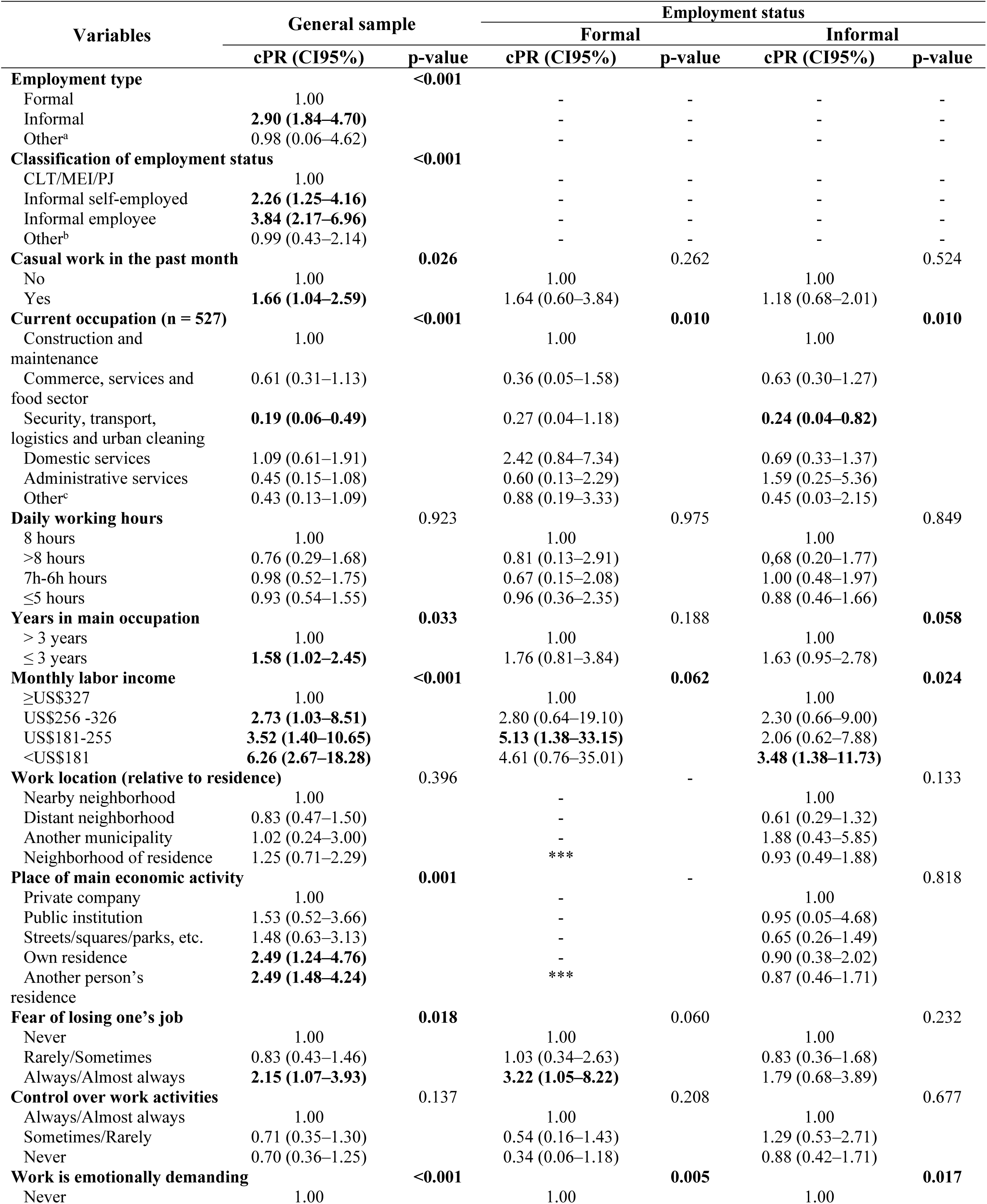
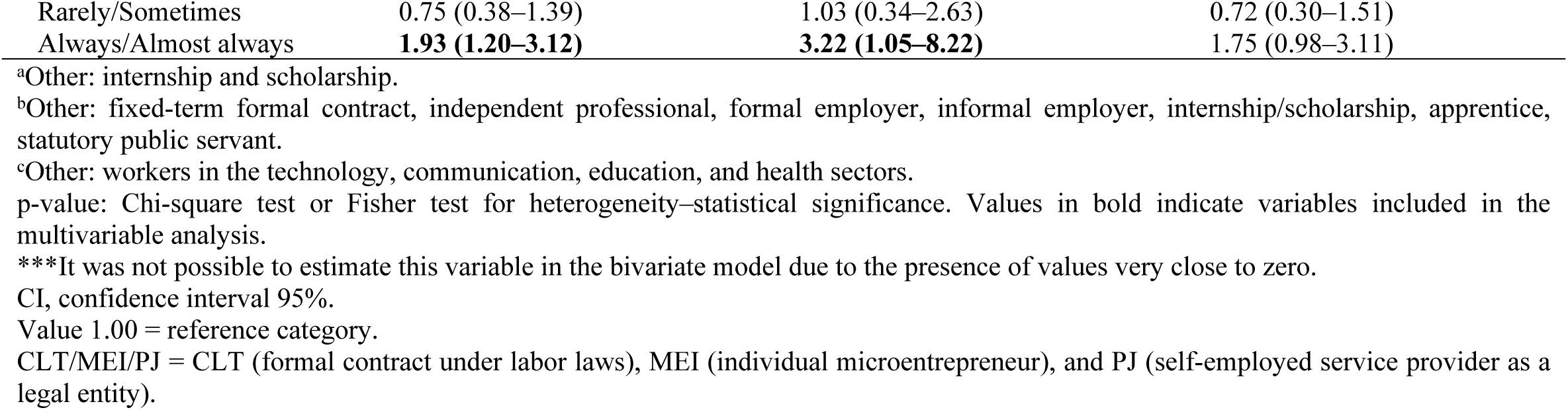
Crude prevalence ratios (cPR) for common mental disorders (CMD) and associated occupational characteristics among workers from urban informal settlements in Salvador, Bahia, Brazil, 2023–2025, by overall sample and employment status.

| Variables | General sample |  | Employment status |  |  |  |
| --- | --- | --- | --- | --- | --- | --- |
|  |  |  | Formal |  | Informal |  |
|  | cPR (CI95%) | p-value | cPR (CI95%) | p-value | cPR (CI95%) | p-value |
| **Employment type** |  | **<0.001** |  |  |  |  |
| Formal | 1.00 |  | - | - | - | - |
| Informal | **2.90 (1.84–4.70)** |  | - | - | - | - |
| Other<sup>a</sup> | 0.98 (0.06–4.62) |  | - | - | - | - |
| **Classification of employment status** |  | **<0.001** |  |  |  |  |
| CLT/MEI/PJ | 1.00 |  | - | - | - | - |
| Informal self-employed | **2.26 (1.25–4.16)** |  | - | - | - | - |
| Informal employee | **3.84 (2.17–6.96)** |  | - | - | - | - |
| Other<sup>b</sup> | 0.99 (0.43–2.14) |  | - | - | - | - |
| **Casual work in the past month** |  | **0.026** |  | 0.262 |  | 0.524 |
| No | 1.00 |  | 1.00 |  | 1.00 |  |
| Yes | **1.66 (1.04–2.59)** |  | 1.64 (0.60–3.84) |  | 1.18 (0.68–2.01) |  |
| **Current occupation (n = 527)** |  | **<0.001** |  | **0.010** |  | **0.010** |
| Construction and maintenance | 1.00 |  | 1.00 |  | 1.00 |  |
| Commerce, services and food sector | 0.61 (0.31–1.13) |  | 0.36 (0.05–1.58) |  | 0.63 (0.30–1.27) |  |
| Security, transport, logistics and urban cleaning | **0.19 (0.06–0.49)** |  | 0.27 (0.04–1.18) |  | **0.24 (0.04–0.82)** |  |
| Domestic services | 1.09 (0.61–1.91) |  | 2.42 (0.84–7.34) |  | 0.69 (0.33–1.37) |  |
| Administrative services | 0.45 (0.15–1.08) |  | 0.60 (0.13–2.29) |  | 1.59 (0.25–5.36) |  |
| Other<sup>c</sup> | 0.43 (0.13–1.09) |  | 0.88 (0.19–3.33) |  | 0.45 (0.03–2.15) |  |
| **Daily working hours** |  | 0.923 |  | 0.975 |  | 0.849 |
| 8 hours | 1.00 |  | 1.00 |  | 1.00 |  |
| >8 hours | 0.76 (0.29–1.68) |  | 0.81 (0.13–2.91) |  | 0.68 (0.20–1.77) |  |
| 7h-6h hours | 0.98 (0.52–1.75) |  | 0.67 (0.15–2.08) |  | 1.00 (0.48–1.97) |  |
| ≤5 hours | 0.93 (0.54–1.55) |  | 0.96 (0.36–2.35) |  | 0.88 (0.46–1.66) |  |
| **Years in main occupation** |  | **0.033** |  | 0.188 |  | **0.058** |
| > 3 years | 1.00 |  | 1.00 |  | 1.00 |  |
| ≤ 3 years | **1.58 (1.02–2.45)** |  | 1.76 (0.81–3.84) |  | 1.63 (0.95–2.78) |  |
| **Monthly labor income** |  | **<0.001** |  | **0.062** |  | **0.024** |
| ≥US\$327 | 1.00 | | 1.00 | | 1.00 | |
| US\$256–326 | **2.73 (1.03–8.51)** | | 2.80 (0.64–19.10) | | 2.30 (0.66–9.00) | |
| US\$181–255 | **3.52 (1.40–10.65)** | | **5.13 (1.38–33.15)** | | 2.06 (0.62–7.88) | |
| <US\$181 | **6.26 (2.67–18.28)** | | 4.61 (0.76–35.01) | | **3.48 (1.38–11.73)** | |
| **Work location (relative to residence)** |  | 0.396 |  | - |  | 0.133 |
| Nearby neighborhood | 1.00 |  | - |  | 1.00 |  |
| Distant neighborhood | 0.83 (0.47–1.50) |  | - |  | 0.61 (0.29–1.32) |  |
| Another municipality | 1.02 (0.24–3.00) |  | - |  | 1.88 (0.43–5.85) |  |
| Neighborhood of residence | 1.25 (0.71–2.29) |  | *** |  | 0.93 (0.49–1.88) |  |
| **Place of main economic activity** |  | **0.001** |  | - |  | 0.818 |
| Private company | 1.00 |  | - |  | 1.00 |  |
| Public institution | 1.53 (0.52–3.66) |  | - |  | 0.95 (0.05–4.68) |  |
| Streets/squares/parks, etc. | 1.48 (0.63–3.13) |  | - |  | 0.65 (0.26–1.49) |  |
| Own residence | **2.49 (1.24–4.76)** |  | - |  | 0.90 (0.38–2.02) |  |
| Another person's residence | **2.49 (1.48–4.24)** |  | *** |  | 0.87 (0.46–1.71) |  |
| **Fear of losing one's job** |  | **0.018** |  | 0.060 |  | 0.232 |
| Never | 1.00 |  | 1.00 |  | 1.00 |  |
| Rarely/Sometimes | 0.83 (0.43–1.46) |  | 1.03 (0.34–2.63) |  | 0.83 (0.36–1.68) |  |
| Always/Almost always | **2.15 (1.07–3.93)** |  | **3.22 (1.05–8.22)** |  | 1.79 (0.68–3.89) |  |
| **Control over work activities** |  | 0.137 |  | 0.208 |  | 0.677 |
| Always/Almost always | 1.00 |  | 1.00 |  | 1.00 |  |
| Sometimes/Rarely | 0.71 (0.35–1.30) |  | 0.54 (0.16–1.43) |  | 1.29 (0.53–2.71) |  |
| Never | 0.70 (0.36–1.25) |  | 0.34 (0.06–1.18) |  | 0.88 (0.42–1.71) |  |
| **Work is emotionally demanding** |  | **<0.001** |  | **0.005** |  | **0.017** |
| Never | 1.00 |  | 1.00 |  | 1.00 |  |

|  |  |  |  |
| --- | --- | --- | --- |
| Rarely/Sometimes | 0.75 (0.38–1.39) | 1.03 (0.34–2.63) | 0.72 (0.30–1.51) |
| Always/Almost always | **1.93 (1.20–3.12)** | **3.22 (1.05–8.22)** | 1.75 (0.98–3.11) |

<sup>a</sup>Other: internship and scholarship.
<sup>b</sup>Other: fixed-term formal contract, independent professional, formal employer, informal employer, internship/scholarship, apprentice, statutory public servant.
<sup>c</sup>Other: workers in the technology, communication, education, and health sectors.
p-value: Chi-square test or Fisher test for heterogeneity–statistical significance. Values in bold indicate variables included in the multivariable analysis.
\*\*\*It was not possible to estimate this variable in the bivariate model due to the presence of values very close to zero.
CI, confidence interval 95%.
Value 1.00 = reference category.
CLT/MEI/PJ = CLT (formal contract under labor laws), MEI (individual microentrepreneur), and PJ (self-employed service provider as a legal entity).

The variables "current occupation" and "emotional demands at work" were excluded from the final multivariable models. The former was excluded because the aggregation of diverse occupations into broad analytical categories resulted in imprecise estimate. The latter was excluded due to the potential for reverse causality. In addition, variables with categories containing zero observations were removed from both crude and final models to avoid model misfit.

### Ethical aspects

The study was approved by the Research Ethics Committee (CAAE 75382123.0.0000.5030; Opinion No. 6.515.557; November 21, 2023). All participants provided written informed consent in accordance with Resolution 466/2012 of the National Health Council. Participants’ data were anonymized to ensure confidentiality. In addition, informational material describing free or low-cost psychosocial and mental health services available in Salvador were provided to participants who, following the interviews scored above the established cutoff point.

## RESULTS

### Sociodemographic characteristics

Between December 2023 and March 2025, 587 workers were interviewed, of whom 332 (56.2%) were formally employed and 242 (41.2%) were informally employed. Overall, 50.9% of participants were female. Although men constituted the majority among formally employed workers (55.7%), women were more prevalent in the informal sector (55.4%). Approximately 29% of participants were aged 40-49 years, whereas only 5.1% aged 60-70 years. Most participants self-identified as Black (59.6%), followed by mixed-race (36.6%), representing about 97% identifying as Black or mixed-race. Regarding educational attainment, 54.9% of participants had ≥12 years of education (completed high school), whereas 45.1% had ≤11 years of education (did not complete high school), this proportion increased to 57.7% among informal workers. Nearly half of respondents were married or in a stable union (48.0%), and 19.1% reported being the primary caregiver for at least a child. Among participants with at least one living child, 35.0% were beneficiaries of the Bolsa Família Program, a Brazilian federal conditional cash transfer program that provides financial support to low-income families contingent on meeting requirements such as school attendance, childhood vaccination, and health monitoring [34]. Only 15.8% of formally employed workers reported receiving the benefit, compared with 45.9% of informal workers (Table 1).

### Occupational characteristics

Regarding occupational profile, the most common employment arrangements were CLT employment (40.7%), informal self-employment (26.3%), and informal wage employment (16.8%). Overall, 23.8% of workers reported performing “casual work” or additional activities to supplement income in the past month. This proportion was higher among informal workers (36.1%) than among formal workers (15.5%). Across the entire sample, the sectors employing the largest proportion of workers were commerce, services, and food (26.0%), followed by civil construction and maintenance (20.7%). Among formally employed workers, the largest share worked in security, transportation, logistics, and urban sanitation (24.0%), whereas among informal workers the most common sector was commerce, services, and food (33.2%) (Table 2; Supplementary Material 1). An 8-hour workday was the most common schedule in the overall sample (43.6%). However, a substantial proportion of workers reported ≤ 5-hour workdays (29.2%), suggesting underemployment. Extended work hours (> 8-hour) were reported by 9.1% of formally employed workers and by nearly one-quarter of informal workers. Regarding income, only 25% of workers reported a monthly income ≥$327, while 46.3% of informal workers reported earnings below < $181 (Table 2).

Regarding occupational characteristics, 64.2% of participants reported having worked in their main occupation for more than three years. In the overall sample, most workers were employed in distant neighborhoods (45.5%) and worked primarily in private companies (50.1%). In contrast, informal workers were more likely to work within their own neighborhood (41.7%) or in another person’s residence (39.7%). With respect to psychosocial characteristics, 5.6% of participants reported always or almost always experiencing conflicts at work, and 6.5% reported always or almost always fearing losing their job. Regarding autonomy in work activities, 64.4% stated that they always or almost always had freedom to perform their tasks, whereas 18.7% reported never having such freedom. Additionally, 27.5% of participants reported that their work was always or almost always emotionally demanding (Table 2).

### Common mental disorders (CMDs)

The overall prevalence of CMD was 14.0% (95% CI: 11.3-17.0), corresponding to 82 cases among the 587 participants who reported symptoms in the previous thirty days. CMD prevalence differed markedly by employment type: 7.8%, among formally employed workers and 22.7% among informal workers, nearly three times higher. Higher prevalences were observed among informal workers with the following sociodemographic characteristics: female (26.1%), aged 18-29 years (34.7%), mixed race (26.5%), 9-11 years of schooling (30.9%), single (25.7%), and those reporting being the primary caregivers for children (32.1%) (Table 1).

Regarding occupational characteristics, informal workers showed the highest prevalence of CMDs (30.5%). When stratified by occupation, higher prevalences were observed among workers in domestic services (25.0%) and workers in commerce, services, and food services (31.5%). Shorter job tenure (≤ 3 years) was also associated with higher CMD prevalence, affecting 18.4% of workers overall and 30.5% of those employed informally. Lower monthly income (<$181) was similarly associated with increased prevalence (24.6%), particularly among informal workers (30.3%). Workplace location was also relevant: higher prevalences were observed among individuals working in their own residence (22.0%) or another person’s residence (22.1%). Regarding psychosocial work factors, frequent fear of job loss (28.9%) and perceiving work as highly emotionally demanding (22.6%) were associated with higher levels of worker distress among participants and across employment types (Table 2).

Tables 3 and 4 present the bivariate models used to guide the selection of variables for the final models. Multivariable models adjusted for sociodemographic and occupational determinants are presented in Table 5. In the overall sample, workers aged 40-49 years showed a 57% lower prevalence of CMD compared with those aged 18-29 years (PR: 0.43; 95% CI 0.19–0.93). Informally employed workers had 2.31 times the prevalence of CMD compared with workers formally employed under the CLT regime (PR: 2.31; 95% CI 1.12-4.83). Workers who reported always or almost always fearing job loss had a 2.32-fold higher prevalence of CMD compared with those who never experienced this fear (PR: 2.32; 95% CI 1.05-4.70). Although not statistically significant, monthly lower labor income <$181 was associated with a higher probability of CMD (PR: 2.46; 95% CI 0.99-7.09) compared with higher income levels (Table 5). In the stratified analysis, among formally employed workers, being female (PR: 5.16; 95% CI 1.65-22.81) and reporting frequent fear of job loss (PR: 3.23; 95% CI 1.00-9.02) were associated with higher CMD prevalence. Among informally employed workers, only monthly income <$181 was associated with increased CMD prevalence (PR: 3.26; 95% CI 1.14-11.77). The wide confidence intervals observed in the stratified models likely reflect the limited number of observations within subgroup. These results should therefore be interpreted with caution, highlighting the need for further studies focusing on workers in marginalized urban communities.

**Table 5.**
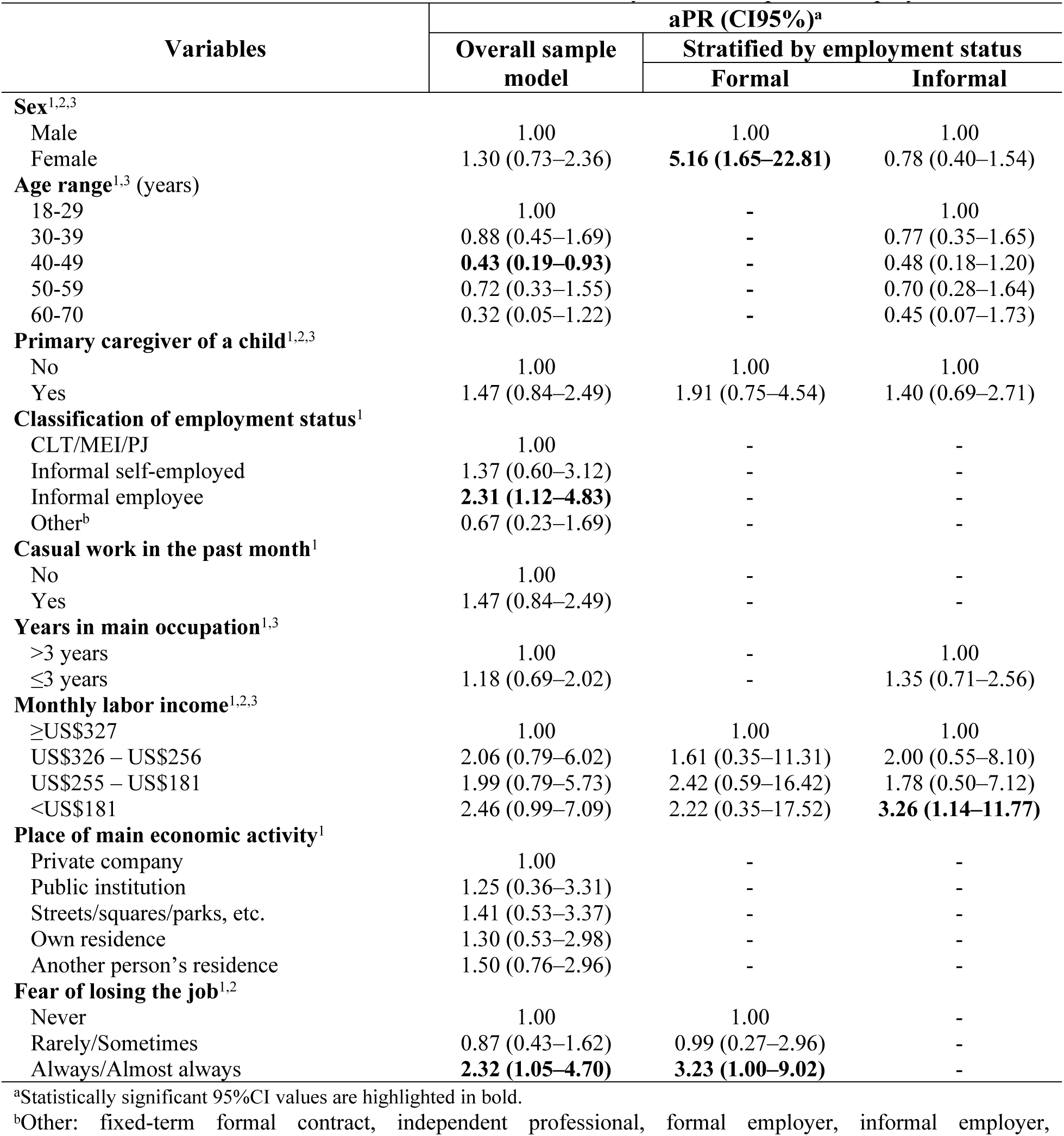
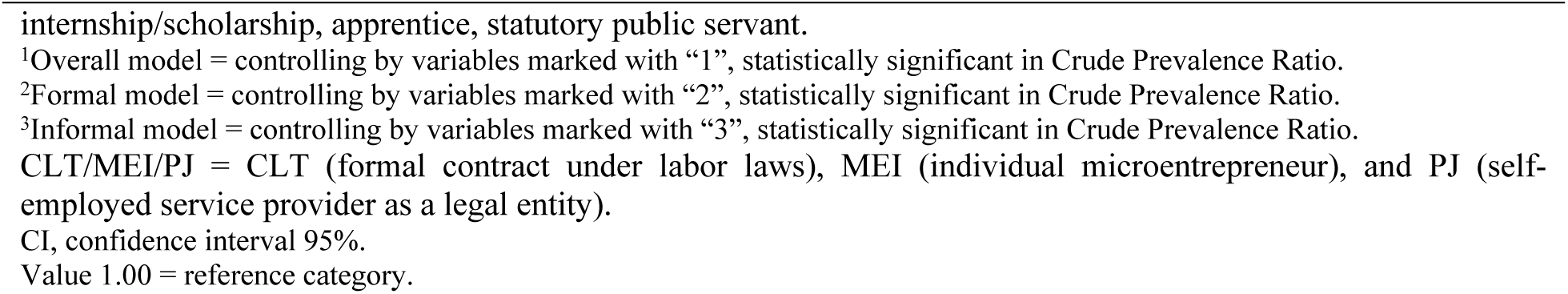
Adjusted prevalence ratio (aPR) of common mental disorders among workers from urban informal settlements in Salvador, Bahia, Brazil, 2023–2025, by overall sample and employment status.

## DISCUSSION

This study aimed to characterize the socio-occupational profile and estimate the prevalence of CMDs and associated determinants among workers residing in urban informal settlements in Salvador, between 2023 and 2025. The overall CMD prevalence was 14.0%, increasing to 22.7% among informal workers. Among participants, CMD was more prevalent among younger workers, those employed informally, and those reporting recurrent fear of job loss. After stratification, formally employed workers presented prevalences below the overall sample average. Among this group, being female and reporting recurrent fear of job loss was associated with higher CMD prevalence. Informal employment was associated with greater psychological distress, with the highest prevalences observed among workers in this group. In the multivariable model, monthly income <$181 was the only factor significantly associated with CMD, although prevalences above the overall sample average were observed across variables examined.

Given the number of limited studies focusing specifically on workers living in urban informal settlements, comparisons were made with research examining mental health of populations with similar social and occupational conditions, such as marginalized urban workers, informal workers, and individual in precarious employment. A meta-analysis of Brazilian studies assessing mental health disorders across various occupational groups reported an average prevalence of approximately 30% among Brazilian workers [35]. In this context, the prevalence observed here is comparable to that reported for bus drivers and conductors (18%), military firefighters (16%), and maritime workers (14%) [35]. Considering the pronounced regional and socioeconomic inequalities in Brazil, comparisons were also made with studies conducted in the Northeast region using the same instrument, although these studies were conducted more than two decades ago. In 2002, a survey of 1,311 urban workers in Feira de Santana, the second largest city in Bahia State, reported a prevalence of mental disorders of 25.2% [36]. Similarly, Ludermir and Lewis [37] analyzed 683 workers in Olinda and found prevalences of 20.7% among formally employed workers, and 35.4% among informal workers, both higher than those observed in the present study [37]. These findings contrast with the initial hypothesis of this study, which assumed that the multiple social determinants affecting workers living in urban communities would result in higher levels of mental distress compared with the broader population of urban workers.

Recent studies conducted in urban informal settlements provide evidence consistent with the findings of the present study and suggest possible hypotheses. Data from the 2019 PNS indicate a lower prevalence of depressive symptoms among favela residents compared with the broader urban population [25]. Similarly, a study conducted in slums in Ibadan, Nigeria, reported a prevalence of CMDs of 14% among female residents, lower that the authors expected [38]. Factors such as strong social network, community cohesion, resilience, and multigenerational cohabitation have been highlighted as important protective elements for the mental health of community residents [25,38–41]. In addition, analyses from the 100 million Brazilians Cohort [42,43] suggest that the expansion of social protection policies over the past 25 years has contributed to improvements in population health and may also influence mental health outcomes among socially vulnerable groups. A notable example is the Bolsa Família conditional cash transfer program, implemented in 2004, which has been associated with a reduced suicide among individuals experiencing socioeconomic vulnerability [44–46]. Together, these findings help contextualize and interpret the results observed in this study.

Two methodological aspects of this study may also have influenced the observed prevalences estimates: the exclusion of unemployed individuals and the choice of the cutoff point for outcome assessment. First, unemployed residents of urban communities were not included in the sample. This population generally exhibits higher prevalences of CMDs than employed population [29,47], which may have contributed to the lower overall prevalence observed. Second, the cutoff point used for the SRQ-20, the instrument used to evaluate CMD, may have affected the results. The SRQ-20 was developed by the WHO to support epidemiological studies and mental health screening in LMICs [48]. In this study, cutoff point of ≥8 positive responses for women and ≥6 for men was adopted, following the validation study by Mari and William [30], which identified difference in the expression of psychological distress and recommended gender-specific thresholds. Although this approach has been used in some Brazilian studies, most national surveys (44.9%) using the SRQ-20 applies a single cutoff point for both sexes [35]. The adoption of gender-specific thresholds in the present study may therefore have contributed to lower prevalences estimates compared with those reported in other studies.

However, several socio-occupational determinants were associated with worse CMDs. In particular, when analyzed separately, the prevalence of CMDs among informal workers was 22.7%, reaching 30.5% among those informally employed by companies or individuals (Table 2). This difference remained statistically significant in the multivariable model, in which these workers had a 2.31-fold higher prevalence of CMD compared with formally employed workers (Table 5). These findings suggest that the nature of informal employment itself contributes to increased psychological distress. Previous studies have consistently reported poorer health outcomes among informal workers, who are often exposed to adverse working conditions, including the absence of social protection, low wages, job insecurity, precarious employment arrangements, higher exposure to violence, chronic stress, and broader socioeconomic vulnerability [11,12,37]. This context has been further intensified in recent years by expansion of the platform economy, labor reforms, and the economic consequences of the COVID-19 pandemic. An important contribution of this study is the differentiation within the heterogeneous group commonly called "informal workers." The results indicate distinct levels of vulnerability between those who work independently in informal activities and those who provide services to employers without a formal employment contract. The latter group faces not only economic instability and the absence of labor protections but also greater exposure to exploitation and violence [49,50]. Recognizing these distinctions is essential for advancing the debate on labor informality and for understanding the multiple layers of vulnerability to mental health problems that coexist within this widespread occupational condition in LMICs.

Informality among study participants warrants attention. This phenomenon is widespread across countries in the Global South. In Brazil, it is particularly concentrated in the North and Northeast regions. Long-standing regional inequalities in income distribution and access to formal employment, especially between the South/Southeast and North/Northeast regions, partly explain the high levels of labor informality observed in Salvador. In Salvador, the informality rate reaches 38%, the highest among Brazilian capitals [15]. The situation is even more pronounced among workers residing in urban informal settlements in Salvador, where 41% are informally employed and 45% have not completed high school. Although Bahia State has the second-largest gross domestic product in the Northeast, political and economic priorities may contribute to the persistence of socioeconomic vulnerability. Since the 1990s, Urban policies in Salvador have emphasized the "entrepreneurialization” of urban communities, prioritizing sectors such as tourism and construction and directing substantial investment toward economic elites [49,51]. For much of the population, this approach only generates temporary and precarious employment opportunities. At the same time, limited investment in social public policies, particularly education and social assistance, may further reinforce structural vulnerabilities and contribute to high levels of labor informality observed in the city.

Another notable finding was the higher prevalence of CMDs among younger workers compared with older workers. Specifically, workers aged 40-49 years showed a 57% lower prevalence of CMDs than those aged 18-29 years (Table 5). This result aligns with recent studies [52,53] suggesting an emerging trend that contrasts with earlier findings [36,37]. Similarly, Corseuil et al. [54] reported that younger workers have historically been more vulnerable during periods of economic recession. Data from the continuous national household sample survey show a marked increase in long-term unemployment among young people between 2013 and 2022, rising from 29.9% (2013) to 38.8% (2019), affecting more than 1.7 million individuals [54]. During the same period, approximately 53% of young workers ventured into the informal labor market. Barriers such as declining availability of formal jobs, low wages, increasing qualification requirements, and the expansion of precarious employment arrangements, including platform-based work and outsourcing, have made it more difficult for younger individuals to establish stable careers. In contrast, older workers tend to experience greater labor market stability [54], which may contribute to better mental health outcomes. These structural labor market challenges may play an important role in the elevated levels of psychological distress observed among young workers.

Workers residing in urban informal settlements who reported a constant fear of losing their jobs were 2.32 times more likely to experience CMDs compared with those who did not report such fear (Table 5). According to ILO [9], precarious employment encompasses both objective dimensions, related to the legal status of the employment relationship, and subjective dimensions, such as perceptions of uncertainty and job insecurity [9]. In this context, Fernandes [50] highlight that short-term contracts, job instability, and erosion of labor protections, key features of precarious employment, can increase workers’ vulnerability to CMD [50]. Evidence from a large cross-national study involving more than 11,000 workers from 21 European Union countries showed that precarious employment arrangements were associated with higher levels of CMDs among both men and women [55]. Similarly a Brazilian study of bus drivers and conductors found that greater job insecurity was associated with higher prevalence of CMDs, sleep disorders, depression, and chronic pain [56].

Another determinant associated with CMD is low labor income. In Bahia, the average monthly salary is approximately $386, the third lowest among Brazil’s 27 federative units, behind only Maranhão and Ceará [57]. In the present study, the average salary among the sample was $294, approximately 24% lower than the state average (data not shown in tables). Moreover, nearly half of the informal workers reported earning <$181 per month (Table 2), underscoring the substantial socioeconomic vulnerability faced by this population. The prevalence of CMD among workers in this lowest income group (<$181) was 26%, reaching 30% among informal workers (Table 4), about twice the prevalence observed in the overall sample. These workers were also 2.46 times more likely to experience CMD compared with those with higher incomes (Table 5). Similar patterns have been observed elsewhere. For example, the ISA-Capital population-based study conducted in São Paulo in 2015 reported prevalences of CMD of approximately 28% among individuals earning up to about $300 per month, a finding similar to that observed in the present study [29]. The WHO notes that poverty and mental illness often form a mutually reinforcing cycle: limited economic resources contribute to psychological distress, while illness may reduce an individual’s productivity, leading to further income loss and increased socioeconomic vulnerability for the household [3].

While the prevalence of CMD among male workers was approximately 9%, below the overall study average, higher prevalences were observed among female workers (19.4%) and among those reported being primary caregivers for children (23.2%). Among formally employed workers, gender emerged as a particularly important determinant of CMD, and women were also disproportionately represented in the informal sector (Table 5). These findings suggest that workers’ mental health is closely intertwined with the social construction of gender roles and the sexual division of labor. A meta-analysis of 174 studies conducted across 63 countries, between 1980 and 2013, reported a consistent gender effect on psychological distress [58]. Women showed higher prevalences of mood and anxiety disorders (19.7%), whereas men presented lower prevalences of mental disorders (14.7%). One key mechanism underlying this disparity is the sexual division of labor, which historically assigns women primary responsibility for household and family care while limiting their access to positions of power and economic opportunity. Despite social progress, this division continues to shape contemporary labor markets, contributing to women’s double or triple work burden, greater employment instability, lower representation in leadership positions, wage disparities, and higher exposure to gender-based violence in the workplace [37,59]. These structural inequalities increase women’s vulnerability to adverse health outcomes, particularly higher prevalence of CMDs.

Racial and ethnic inequalities further shape the distribution of socioeconomic vulnerability in Brazil. According to the report of Social Inequalities by Color or Race in Brazil [16], labor market participation differs substantially across racial groups. In 2021, 43% of Black individuals and 47% of mixed-race individuals were employed in the informal sector, compared with 32% of white individuals. In contrast, managerial positions were disproportionately occupied by White workers (69%), while only 29% were held by Black individuals. Historically, precarious employment has been concentrated among socially marginalized groups, including Black and mixed-race individuals, women, younger and older persons, and those residing in rural areas or marginalized urban areas [50,60]. Consistent with this pattern, 96% of workers residing in urban communities in Salvador identified as Black or mixed-race. The unequal distribution of economic opportunities across racial groups is deeply rooted in Brazil’s colonial and slaveholding past, during which racial hierarchies were institutionalized to justify the exploitation of Indigenous and African populations and the expropriation of their land, labor, and resources [17]. These historical processes contributed to the accumulation of wealth among white elites and established structural inequalities that persist today. As a result, racial and socioeconomic disparities remain embedded in contemporary labor markets, often intensified by neoliberal economic dynamics. In this context, Black and mixed-race individuals continue to disproportionately occupy precarious jobs in major urban centers. Milton Santos [19] conceptualized this phenomenon as “perverse globalization”, in which economic, technological, and political systems operate primarily to benefit a privileged minority while reproducing poverty and inequality for the majority. Such structural dynamics help explain the persistence of socioeconomic and racial inequalities observed among the sample [19].

This study has some limitations that should be considered when interpreting its findings. First, the cross-sectional design precludes causal inference and raises the possibility of reverse causality. Although temporal variables were included to help contextualize potential relationships between exposure and CMDs, casual interpretations remain limited. Second, the survey was conducted in only five of the 262 urban informal settlements in Salvador, which may limit the generalizability of the findings. Nevertheless, the sociodemographic characteristics observed in this study are similar to those reported for favelas in Bahia, which show a higher proportion of women (53.4%) and Black residents (39.2%), as well as a relatively young population structure, with only 12.7% of residents aged 60 years or older [20]. Third, the SRQ-20 is a screening instrument and therefore does not allow for clinical diagnosis or differentiation between specific mental disorders, such as anxiety or depressive disorders. However, the SRQ-20 is widely used in epidemiological research and has strong comparability across studies, which facilitates its application in socioeconomically vulnerable populations. Additionally, the possibility of response bias cannot be ruled out, given the sensitive nature of mental health reporting. Another potential limitation relates to the classification of occupations. Due to the substantial heterogeneity of reported jobs, occupations had to be grouped into broader analytical categories, which may have obscured important variations within occupational groups. Thus, standardized occupational classification systems such as the BCO and CNAE could not be applied for comparison. Finally, the relatively small number of observations within the stratified groups of formal and informal workers might have reduced the statistical power and precision of the multivariable analyses, resulting in wider confidence intervals. Future studies with larger and more diverse samples of workers living in marginalized urban communities are therefore needed to further examine the determinants of CMD in these settings.

Despite these limitations, several important contributions of this study should be highlighted. To the authors’ knowledge, this is the first examination of socio-occupational characteristic and prevalence of CMDs among workers residing in urban informal settlements. In this context, the study provides an important descriptive contribution to understanding mental health in a population that has been largely overlooked in occupational health research. These findings should contribute to strengthening public policies related to psychosocial care, professional training, racial reparation, and income distribution for workers residing in urban informal settlements across Brazil. The study also represents a pioneering contribution to the field of occupational and mental health and to the advancement of epidemiological research. Furthermore, the exploration of different forms of informal employments should help to highlight important differences within the heterogonous group of workers commonly referred to as “informal”. This distinction remains insufficiently addressed in the literature and deserves further evaluation in future studies. Ultimately, these results and reflections should contribute to improving the lives of the study population, who are, among the main victims of the social and racial inequalities that have historically affected the country.

## CONCLUSIONS

In this sample of workers residing in urban informal settlements in Salvador, Bahia, Brazil, the prevalence of CMDs was 14.0%, reaching 22.7% among those employed in the informal sector. In the overall sample, informal employment and persistent fear of job loss were associated with higher levels of mental health problems, whereas workers aged 40-49 years presented lower prevalence compared with younger workers. Among formally employed workers, female gender and persistent job insecurity were associated with a greater risk of CMDs. Among informal workers, the employment relationship itself, particularly when combined with very low income, was associated with increased mental health vulnerability. These findings contribute to increasing the visibility of a population that remains largely overlooked in occupational health research. They also highlight the importance of policies aimed at improving employment stability, income security, and access to social protection, as well as broader initiatives to address the structural determinants affecting the mental health and well-being of the population residing in urban informal settlements.

## Acknowledgements

The authors are grateful to all the communities that participated in this study. We also thank the community leaders and participants who supported our work and the research team.

## Availability of data and materials

The datasets used and/or analyzed during the current study cannot be shared publicly because of personal information of participants in the survey, at the individual and household level. Researchers who wish to access the data can contact Dr. Mitermayer Galvão Reis, Principal Investigator, Oswaldo Cruz Foundation or data manager at the Oswaldo Cruz Foundation, Dr. Nivison Nery Junior.

All codes used during this study are available in Zenodo under Creative Commons 4.0 license, accessible through https://doi.org/10.5281/zenodo.19228099.

